# Global convergence of COVID-19 basic reproduction number and estimation from early-time SIR dynamics

**DOI:** 10.1101/2020.04.10.20060954

**Authors:** Gabriel G. Katul, Assaad Mrad, Sara Bonetti, Gabriele Manoli, Anthony J. Parolari

## Abstract

The SIR (‘susceptible-infectious-recovered’) formulation is used to uncover the generic spread mechanisms observed by COVID-19 dynamics globally, especially in the early phases of infectious spread. During this early period, potential controls were not effectively put in place or enforced in many countries. Hence, the early phases of COVID-19 spread in countries where controls were weak offer a unique perspective on the ensemble-behavior of COVID-19 basic reproduction number *R*_*o*_. The work here shows that there is global convergence (i.e. across many nations) to an uncontrolled *R*_*o*_ = 4.5 that describes the early time spread of COVID-19. This value is in agreement with independent estimates from other sources reviewed here and adds to the growing consensus that the early estimate of *R*_*o*_ = 2.2 adopted by the World Health Organization is low. A reconciliation between power-law and exponential growth predictions is also featured within the confines of the SIR formulation. Implications for evaluating potential control strategies from this uncontrolled *R*_*o*_ are briefly discussed in the context of the maximum possible infected fraction of the population (needed for assessing health care capacity) and mortality (especially in the USA given diverging projections). Model results indicate that if intervention measures still result in *R*_*o*_ *>* 2.7 within 49 days after first infection, intervention is unlikely to be effective in general for COVID-19. Current optimistic projections place mortality figures in the USA in the range of 100,000 fatalities. For fatalities to be confined to 100,000 requires a reduction in *R*_*o*_ from 4.5 to 2.7 within 17 days of first infection assuming a mortality rate of 3.4%.

## Introduction

A heated dispute about the effectiveness versus risk of smallpox inoculation was playing out in eighteenth-century France, which was to launch the use of mathematical models in epidemiology. This dispute moved inoculation from the domain of philosophy, religion, and disjointed trials plagued by high uncertainty into a debate about mathematical models - put forth by Daniel Bernoulli (in 1766) and Jean-Baptiste le Rond D’Alembert (in 1761), both dealing with competing risks of death and interpretation of trials [1]. Since then, the mathematical description of infectious diseases continues to draw significant attention from researchers and practitioners in governments and health agencies alike. Even news agencies are now seeking out explanations to models so as to offer advice and clarity to their audiences during the (near-continuous) coverage of the spread of COVID-19 [2]. The prospect of using mathematical models in conjunction with data is succinctly summarized by the Nobel laureate Ronald Ross, whose 1916 abstract [3] enlightens the role of mathematics in epidemiology today. A quotation from this abstract below, which foreshadows the requirements and challenges for mathematical models to describe emerging epidemics such as COVID-19 [4, 5], needs no further elaboration:

> *It is somewhat surprising that so little mathematical work should have been done on the subject of epidemics, and, indeed, on the distribution of diseases in general. Not only is the theme of immediate importance to humanity, but it is one which is fundamentally connected with numbers, while vast masses of statistics have long been awaiting proper examination. But, more than this, many and indeed the principal problems of epidemiology on which preventive measures largely depend, such as the rate of infection, the frequency of outbreaks, and the loss of immunity, can scarcely ever be resolved by any other methods than those of mathematical analysis*.

The classic susceptible-infectious-recovered (SIR) paradigm, initiated in the late 1920s [6], now provides a mathematical framework that describes the core transmission dynamics of a range of human diseases [7–12], including COVID-19 [13]. A key parameter in the SIR paradigm is the basic reproduction number (*R*_*o*_). The *R*_*o*_ is defined by the average number of secondary cases arising from a typical primary case in an entirely susceptible population of size *S*_*o*_ [14–16]. The usefulness of *R*_*o*_ and uncertainty in its estimation are not a subject of debate, as reviewed elsewhere [17], and therefore are not further discussed here.

In the analysis herein, the SIR model is used to uncover generic spread mechanisms observed by COVID-19 dynamics globally, especially in the early phases of infectious spread. During this early period, potential controls were not effectively put in place or enforced in many countries around the world despite early warning signals from China, Iran, and later on, Italy. Hence, the early phases of COVID-19 spread in many countries where controls were weak offer a unique perspective on the ensemble-behavior of COVID-19 *R*_*o*_. The analysis shows that there is global convergence (i.e. across many nations) to an uncontrolled *R*_*o*_ = 4.5 for COVID-19 describing early times spread. The implications for evaluating potential control strategies from this reference *R*_*o*_ are briefly discussed in the context of mortality and maximum infections.

## Theory

### Definitions and Nomenclature

Mathematical models of disease spread assume that a population within a compartment (e.g., city, region, country) can be subdivided into a set of distinct classes [11]. The SIR model classifies individuals in the compartment as one of three classes: susceptible (*S*), infectious (*I*), and recovered or removed (*R*). Infectious individuals spread the disease to susceptible and remain in the infectious class for a given period of time known as the infectious period before moving into the recovered (or removed) class. Individuals in the recovered class are assumed to be immune for an extended period (or removed from the population). For the total population *N* = *S* + *I* + *R*, the dynamical system describing the SIR equations are given as

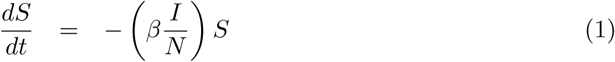

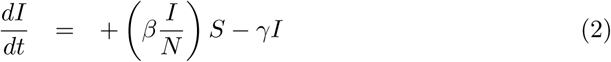

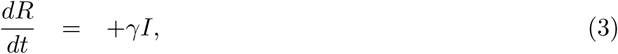

where *λ*(*I*) = *β*(*I/N*) is known as the force of infection and coefficients *β* and *γ* must be externally supplied. Moreover, this system requires the specification of 3 initial conditions *S*(0), *I*(0), and *R*(0). For COVID-19, it is assumed that *R*(0) = 0 and *I*(0) *<< S*(0). For the initial conditions selected here, *N* = *S*(0) + *I*(0) + *R*(0) *≈ S*(0), which is labeled *S*_*o*_ for consistency with the SIR literature. The basis of the latter assumption is that the number of deceased individuals is *<< N*. The dynamical system in equation (3) has only one equilibrium point: *I* = 0 for any *S* and *R*, which is a disease-free stable equilibrium. The SIR model makes a number of assumptions, including a closed system with no changes in natural births or natural deaths occurring during the short-lived outbreak. The infection is assumed to have negligible latent period so that an individual becomes infectious when infected. Disease transmission occurs through individual-to-individual contact directly (skin-to-skin), indirectly (skin-infected surfaces), or airborne (pathogens transmitted through air by small particles after coughing or sneezing). Recovering from infection is also assumed to confer long-term immunity, yet to be verified for COVID-19. The most objectionable assumption in SIR dynamics is the use of the mass-action principle. As with all compartment models, mass action assumes that the rate of encounter between *I* and *S* is proportional to their product. For this assumption to hold, it requires that members of *I* and *S* be uniformly distributed in the space of the compartment [18]. Individuals – unlike molecules in an ideal solution within a closed container – do not mix homogeneously. At minimum, the use of the mass action principle serves as a reference model to compare more detailed mechanisms or explore data.

The parameters *γ* and *β* encode the main properties of the epidemics and the population response to it. The *γ* = 1*/D* is generally interpreted as the inverse of the mean recovery time *D*. The *D* varies with the nature of the disease and the recovery from it which depends on the medical facilities and resources available. For COVID-19, the best information on the speed of recovery comes from a World Health Organization study examining more than 55000 cases in China [19]. They found that for mild illness, the time from the onset of symptoms to natural recovery is, on average, 14 days. This estimate was also supported in other published studies (e.g., [20]), though as much as 6-8 weeks were recorded for severe infections. Because *I* is dominated by mild cases thus far, *D* = 14 d is selected here.

With this assumption, the remaining model parameter *β* must be determined empirically or from separate studies. The *β* reflects the multiplicative effect of two factors: (1) the transmissibility of the infectious disease (= *T*_*r*_) or the probability of disease transmission after an encounter between a susceptible and an infected and (2) the number of contacts per unit time *k* each infected individual has with susceptibles. Hence, *β* = *k T*_*r*_. Factors such as hand-washing and sanitizing reduce *T*_*r*_ whereas social distancing, self-isolation, and closure of public spaces reduce *k*. It is evident that *dI/dt* will be positive (outbreak) or negative (epidemic contained) depending on the sign of (*β*(*S/S*_*o*_) *− γ*), which is one of the main reasons the basic reproduction number is sought.

### The basic reproduction number

As earlier stated, the average rate of recovery is set to *γ* = 1*/D*. Given the value of *D* (in days), the probability that an individual remains infected in an infinitesimal time period *δτ* is 1 *− γ*(*δτ*). Therefore, the probability that this individual remains infected for an amount of time *τ* is lim_*δτ→* 0_(1 *− γδτ*)^*τ/*(*δτ*)^ = exp (*− γτ*). In other words, *τ*, the time that an infected individual remains infected, is exponentially distributed with an average of *D* = 1*/γ*.

In a compartmental model such as the SIR, every individual is susceptible and the average number of susceptibles that encounter an infected individual over a period *τ* is simply *β τ*. It follows that the average number of new infections caused by an infected individual, which is the basic reproduction number *R*_*o*_, is given by [21]

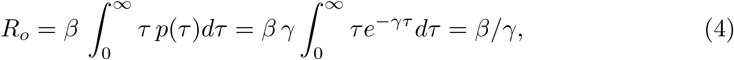

where the *γ* after the second equality is to normalize *p*(*τ*).

While the compartmental SIR model, in use here, assumes an exponentially distributed recovery time and a heavily peaked distribution of encounters with all *S*_*o*_ individuals, the Poisson random graph SIR model assumes a heavily peaked recovery time of *D* = 1*/γ* and a Poisson distributed number of encounters for each individual [22], both more realistic assumptions. However, it can be shown that the dynamics of a discrete-time SIR compartmental model (Reed-Frost model) and the SIR on a Poisson random graph are the same [23]. Because of the aforementioned correspondence, the use of the tractable compartmental SIR model as a proxy to the more complex social network SIR model may be justified here.

### Early-times dynamics of the SIR system

An illustration of the canonical SIR dynamics during an epidemic is shown in Figure 1, where the SIR dynamics is solved here for *S*(*t*), *I*(*t*), and *R*(*t*) when setting *R*_*o*_ = 4.5, *γ* = (1*/*14) d^*−* 1^ and *S*(0) = 100, 000. For small *γt* (dimensionless time), the fraction of susceptibles *S*(*t*)*/S*_*o*_ does not deviate appreciably from unity as seen from Figure 1. For such early times, *I*(*t*) can be made non-dimensional by *S*_*o*_ and decoupled from *S*(*t*) using the approximation

**Fig 1.**
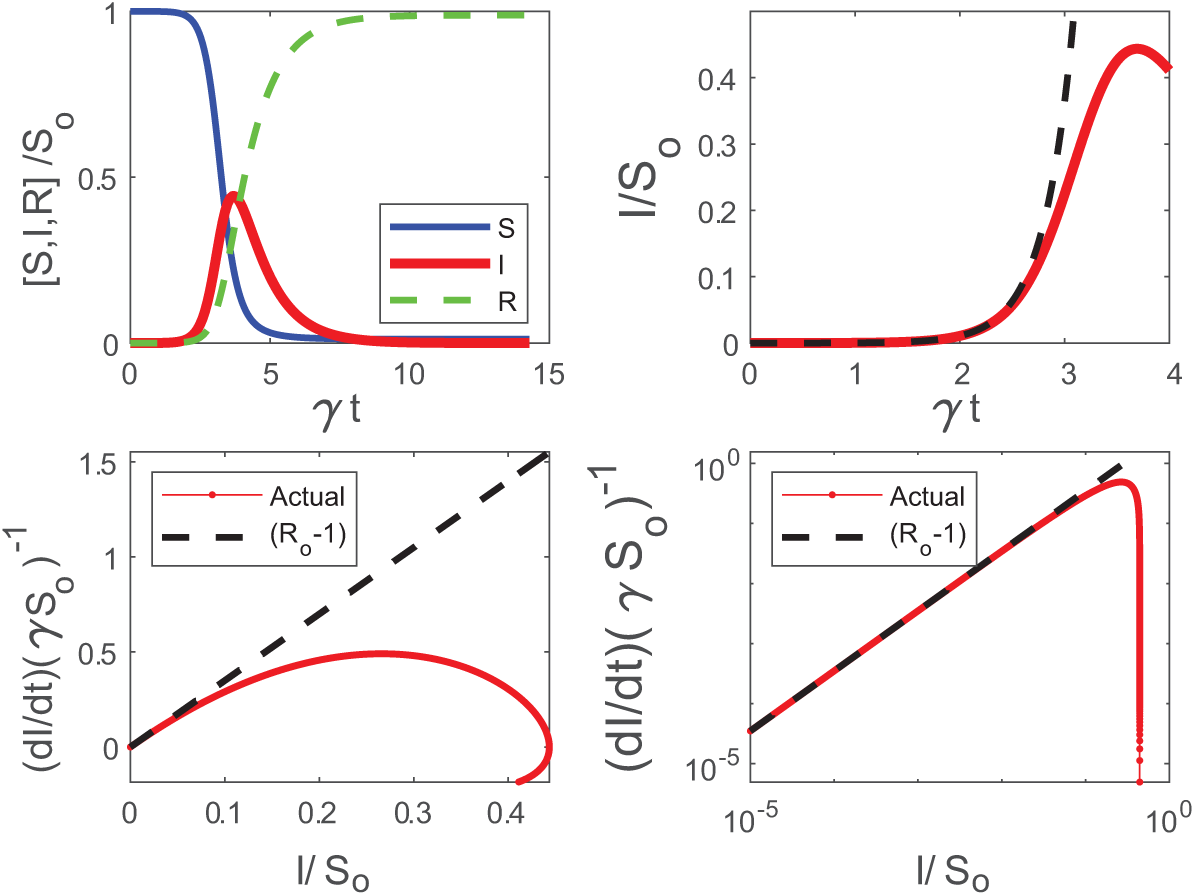
Phase space and temporal trends of the SIR model. Top left: *S*(*t*), *I*(*t*), *R*(*t*) normalized by *S*_*o*_ as a function of dimensionless time *γt* with *S*_*o*_ = 100, 000, *γ* = (1*/*14) d^*−* 1^, and *R*_*o*_ = 4.5. Top right: *i* = *I/S*_*o*_ in dimensionless time *γt* for early times *γt <* 1 revealing strictly exponential growth (dashed) and deviations from exponential (SIR solution). Bottom: (*S*_*o*_*γ*)^*−* 1^*dI/dt* with *I*(*t*)*/S*_*o*_ in linear (left) and double-log (right) representations. The dashed line is (*R*_*o*_*−* 1) where *R*_*o*_ = 4.5. Declines from the dashed line reflect the incipient point where *I*(*t*) deviates appreciably from exponential growth. Note how the early-time slope (*R*_*o*_*−* 1) is emphasized in the double-log representation.

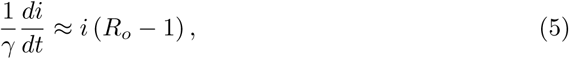

where *i* = *I/S*_*o*_ is the dimensionless fraction of infected individuals and *R*_*o*_ = *β/γ* as before. When *R*_*o*_ *>* 1, *di/dt >* 0 leading to an epidemic or, conversely, a containment of the disease. The solution of equation (5) is an exponential function *I*(*t*)*/I*(0) = exp [(*R*_*o*_ *−* 1) *γt*] also shown in Figure 1.

The *R*_*o*_ may be determined by regressing log(*i*) against *t*, and the slope of this regression determines *R*_*o*_ when *γ* can be separately estimated. More sophisticated fitting procedures can also be conducted on sampled *I*(*t*) versus *t*. A major limitation to this exercise is that *I*(*t*) at early times, often determined from reported confirmed cases, is uncertain and depends on testing frequency that may vary in time as *I* increases. An alternative is to regress *di/dt* upon *i* at early times to detect the highest slope, which can then be used to infer *R*_*o*_. This approach is also featured in Figure 1, which illustrates that the SIR dynamics exhibit rapid deviations from a linear *di/dt* with *i* set by early times thereby underestimating *R*_*o*_ (for a given *γ*). Evidently, inference of *R*_*o*_ requires estimates of early time slope, which cannot be easily detected in practice.

A non-conventional approach is to present confirmed infection data using a double-log representation of *di/dt* versus *i*, which is also featured in Figure 1. This presentation has a number of advantages and limitations in the analysis of COVID-19 discussed elsewhere [24]. The main advantage is that the early time slope (= *R*_*o*_*−* 1) persists over much of the graph. A significant decline in *di/dt* is also required before ‘registering’ a drop in such a representation. This insensitivity to moderate declines in *di/dt* from its initial value may be advantageous in *R*_*o*_ estimates. The other main limitation, which is inherent to all such analyses, is shifts in testing frequency at high *i*, and thus the increase in confirmed cases due to expanded testing. It is to be noted that a log-log representation will be more robust to these shifts, because the overall graph will be biased by the initial slope prior to the initialization of expanded testing. Such bias should lead to increases in *di/dt* versus *i*, not declines from the initial slope (*R*_*o*_ *−* 1) that can be detected. As later shown, such an increase has been noted in several data sets.

With this representation, it is now shown that initial inaction to COVID-19 across many countries around the globe allowed an ensemble estimate of the uncontrolled *R*_*o*_. Because *R*_*o*_ is likely to be at maximum when no action to COVID-19 are implemented early on, a maximum theoretical ‘boundary-line’ can then be derived to describe the spread of COVID-19 for large *S*_*o*_ (on log-log representation). This boundary-line analysis can then be used as a logical reference to assess whether measures to reduce *β* are effective.

## Results and Discussion

### Estimating an early-time *R*_*o*_

The same log-log scheme featured in Figure 1 is now applied to the global data set supplied by the European Center for Disease Prevention and Control (ECDPC). The data source provides daily confirmed infections *I*(*t*) and deaths reported for each country. The population of each country, used to estimate *S*_*o*_ (i.e. all members are susceptible), was obtained from the 2018 United Nations census and provided as part of the ECDPC data base. While daily data are supplied, not all countries report consistently on a daily *I*(*t*). For this reason, daily data on infections were smoothed with a 7 day block-average and *dI/dt* was estimated from the smoothed data. The results show a global convergence to *R*_*o*_ = 4.5 from early time-analysis in Figure 2.

**Fig 2.**
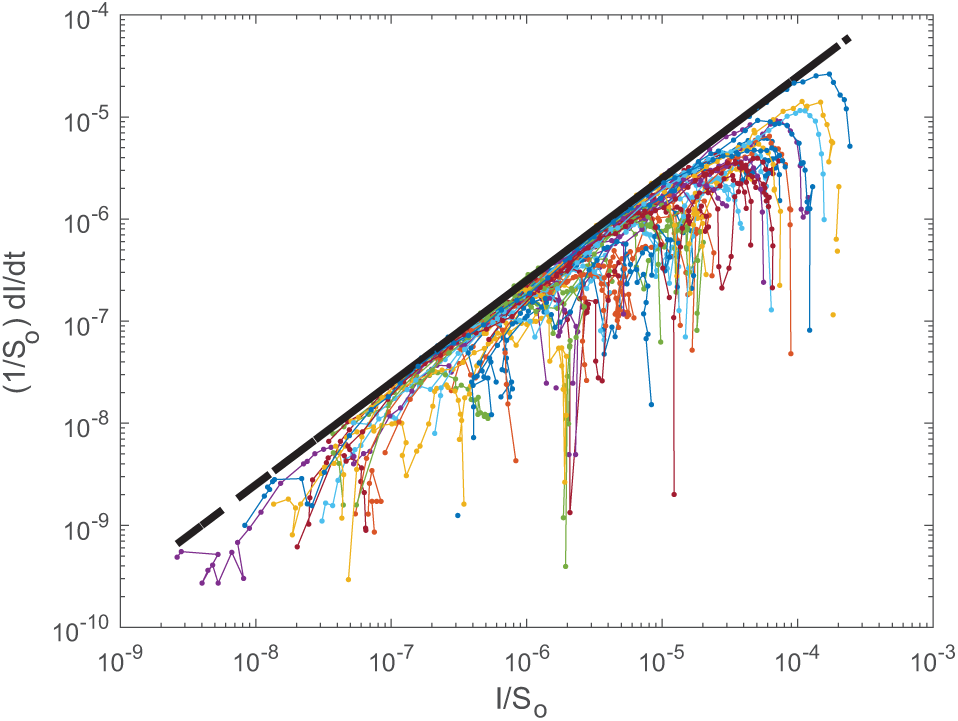
Comparison between (1*/S*_*o*_)(*dI/dt*) and *I/S*_*o*_ for 57 countries. The dashed line is (*R*_*o*_ *−* 1)*γ*, where *R*_*o*_ = 4.5, and *γ* = (1*/*14)*d*^*−* 1^. Negative deviations from the dashed line reflect deviations from exponential.

Examples for specific countries are also featured in Figure 3 illustrating the same early slope patterns. Mindful of all the pitfalls in determining *R*_*o*_ [17], the global estimate here of *R*_*o*_ = 4.5 is roughly commensurate with other entirely independent estimates for COVID-19. The most recent update from the China study suggests an *R*_*o*_ = 4.1 [25] whereas for France, the most recent estimate for early times is *R*_*o*_ = 4.9 [26]. The initially reported and the much cited *R*_*o*_ = 2.2 value [4] from Wuhan, China appears to be low [27]. A more elaborate estimate of *R*_*o*_ based on case reports, incubation periods, high-resolution real-time human travel data, infection data combined with agent-based mathematical models result in *R*_*o*_ = 4.7 *−* 6.6 [27]. Other studies report values between 3.3 and 6.6 [28]. It must be emphasized that the *R*_*o*_ determined here reflects ‘country-scale’ early times assuming the entire country population to be *S*_*o*_, *γ* = (1*/*14)*d*^*−* 1^ and does not accommodate any early measures enacted to reduce *β* or increase *γ*, which were undertaken in China [13].

**Fig 3.**
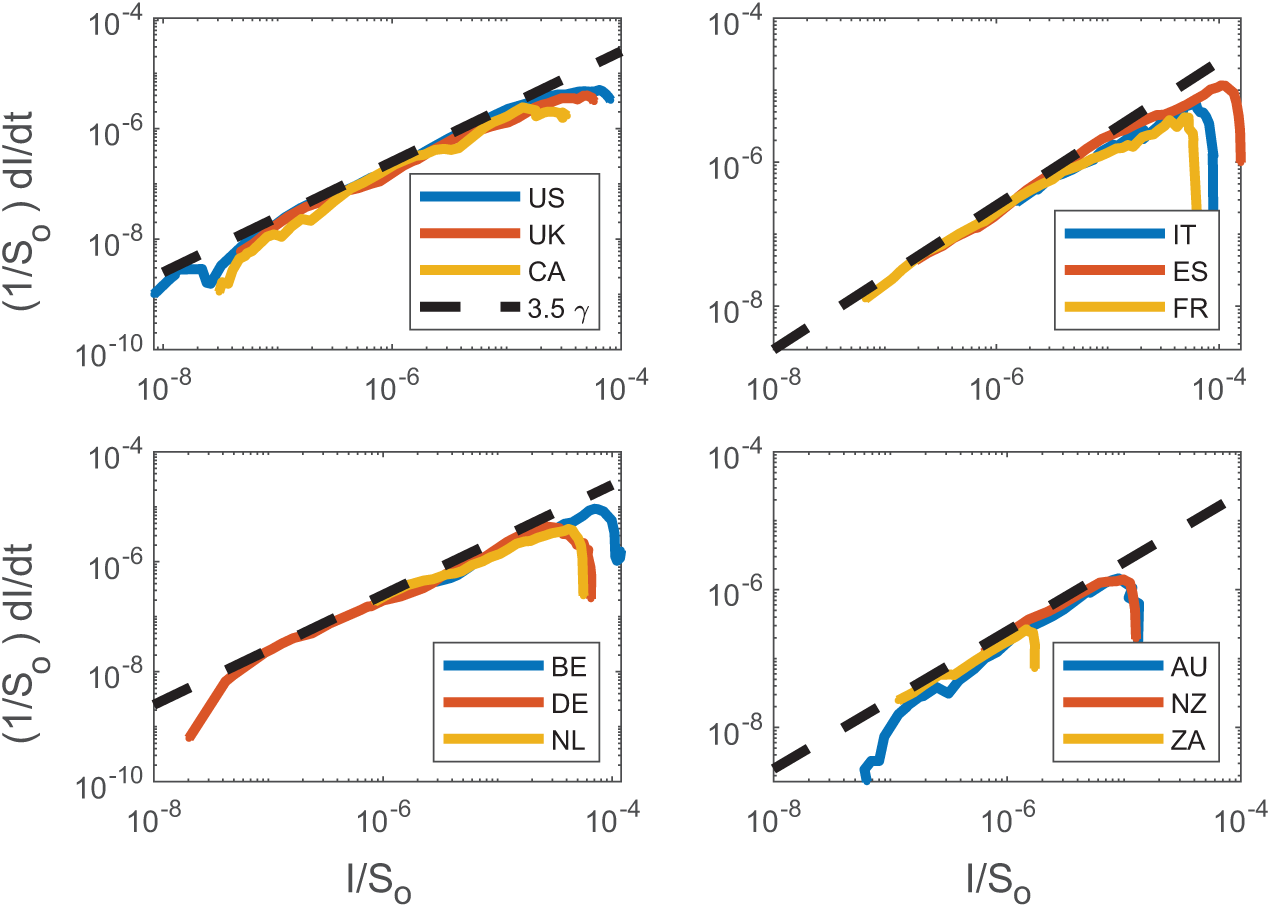
Same as Figure 2 but for sample countries. Top left: the United States of America (US), the United Kingdom (UK), and Canada (CA); bottom left: Belgium (BE), Germany (DE), the Netherlands (NL); top right: Italy (IT), Spain (ES), and France (FR); bottom right: Australia (AU), New Zealand (NZ), and South Africa (ZA).

### Sub-national dynamics and interventions

The same analysis performed for World countries is now applied at a sub-national level, considering Upper Tier Local Authorities (UTLAs) in the UK and provinces in Italy (Fig. 4). Results show a higher variability than country-level data (as expected) but the theoretical ‘boundary-line’ of *R*_*o*_ = 4.5 is shown to hold also at finer spatial scales.

**Fig 4.**
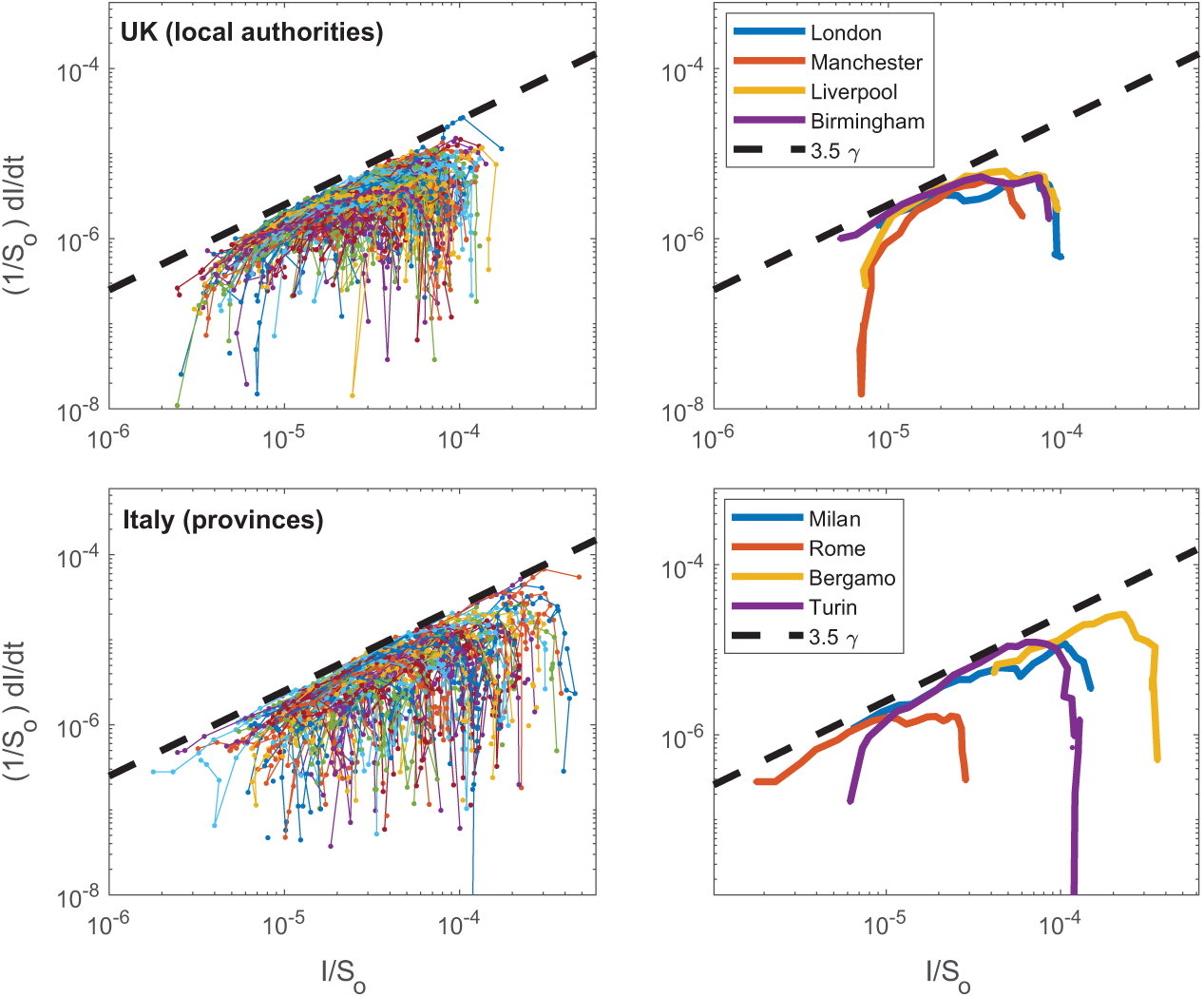
Same as Figure 2 but for sample UTLAs in the UK (top left) and provinces in Italy (bottom left). Selected UTLAs and provinces are shown in the top right and bottom right panels, respectively.

Cases reported at the beginning of April, demonstrate that UK regions are at an early phase of the epidemics (with more ramp-up in testing as later discussed), while Italian provinces are approaching the peak of infections due to strict interventions put in place by national authorities.

To consider the impact of interventions - which have direct effects on local scale dynamics [13] - the SIR model can be solved with a time-varying *R*_*o*_ that decreases from the initial uncontrolled value of 4.5 to, e.g. *R*_*o*_ = 1.1 after 2*/γ* days. To this purpose, the *R*_*o*_ is expressed as a logistic function

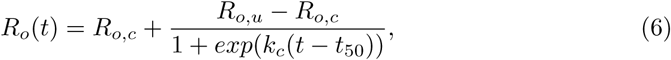

where *R*_*o,c*_ and *R*_*o,u*_ are the ‘controlled’ and ‘uncontrolled’ values of *R*_*o*_ (set to 4.5), *k*_*c*_ is the steepness of the intervention curve and *t*_50_ is the time when *R*_*o*_(*t*) = (*R*_*o,c*_ + *R*_*o,u*_)*/*2. Model results accounting for different intervention scenarios (Fig. 5) resemble the trends observed in the Italian provinces with the timing and magnitude of *R*_*o*_ reductions shifting the linear relation down and decreasing the maximum fraction of infected individuals. Such jumps are smoothed over at the national level where a clear deviation from exponential is observed (Fig. 3).

**Fig 5.**
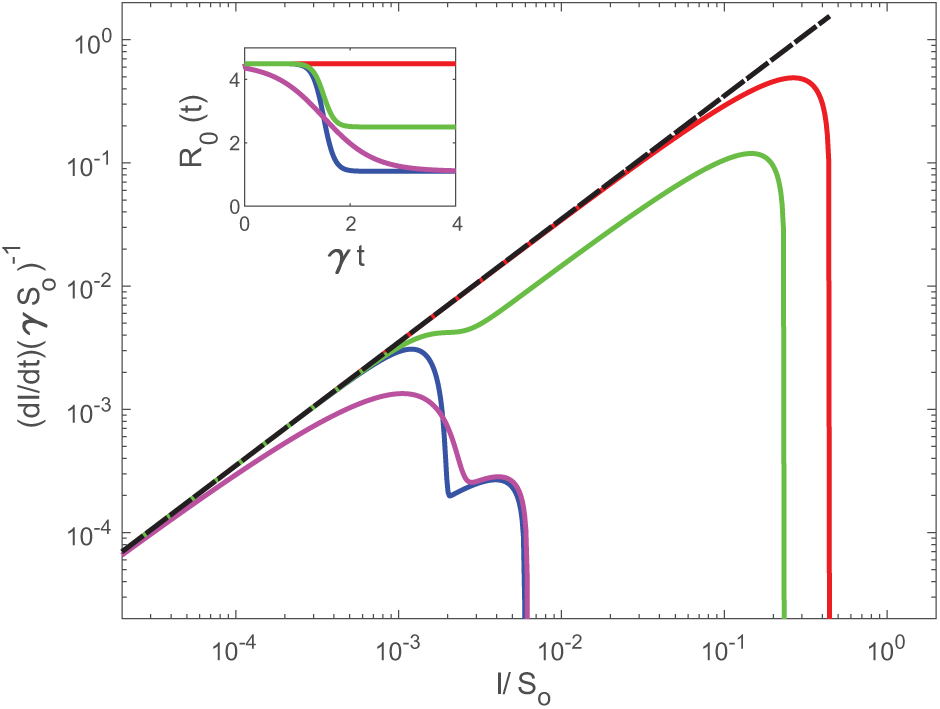
Modeled (1*/S*_*o*_)(*dI/dt*) as a function of *I/S*_*o*_ when considering a time-varying *R*_*o*_(*t*). Five scenarios are illustrated (inset): no intervention (red) with *R*_*o*_ = 4.5 set to its uncontrolled value, *R*_*o,c*_ = 1.1 (epidemic near containment) and *k*_*c*_ = 0.7 (blue), *R*_*o,c*_ = 1.1 and *k*_*c*_ = 0.15 (magenta), *R*_*o,c*_ = 2.5 (typical of countries with strong initial intervention) and *k*_*c*_ = 0.7 (green). The other parameters of the logistic functions are *R*_*o,u*_ = 4.5 and *t*_50_ = 1.5*/γ*.

### An alternative hypothesis: power-law vs exponential

Whether these results are suggestive of a global convergence to an uncontrolled *R*_*o*_ = 4.5 or to some other dimensionless property must not be overlooked. A linear relation on a log-log representation may also be indicative of power-law solutions at early times, already documented in a number of studies for COVID-19 [29, 30]. In fact, published analysis of infection data from the top 25 affected countries reveals approximate power-law behavior of the form *I∼t*^*a*^ (or log(*i*) = *a* log(*t*) + *b*) with two different growth patterns [29]: steady power law growth with moderate scaling exponents (i.e., *a* =3-5) or explosive power law growth with dramatic scaling exponents (i.e., *a* =8-11).

Within the confines of the SIR dynamical system framework here, we ask: what are the necessary modifications to obtain power-law solutions at early times? Such a solution, while not unique, may be possible by revising the force of infection as *λ*(*I*) = *β*(*I/N*)^*m*^. The original SIR model is recovered when *m*=1. For this non-linear force of infection, the SIR system becomes

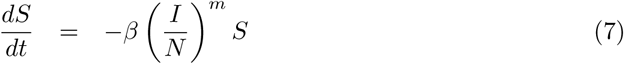

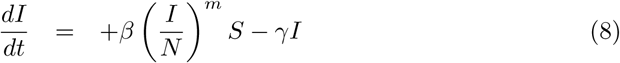

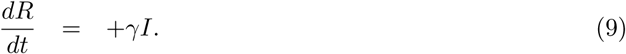

This revision ensures that the total population maintains its constant value *N ≈ S*_*o*_ here. The early times dynamics (i.e. *S*(*t*) *≈ S*_*o*_) for the non-dimensional infection compartment *i* = *I/S*_*o*_ are now governed by

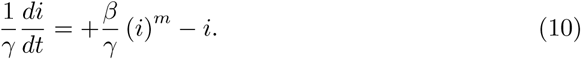

When *m <* 1, maintaining a definition of *R*_*o*_ = *β/γ >* 1 (epidemic), and noting that *i <<* 1, the first term on the right-hand side of equation (10) is much larger than the second term. In fact, to obtain a maximum exponent enveloping the early-time relation between *di/dt* and *i*, the linear term can be dropped so that *di/dt≈ βi*^*m*^ (only a growth phase). On a log-log representation, log(*di/dt*) = *m* log(*i*) + log(*β*). A constant slope such as those featured in Figures 2 and 3 may simply be estimates of *m* instead of *R*_*o*_. The initial conjecture is that a power-law solution emerges from the modified SIR dynamics when *m <* 1. However, the slope here (= 3.5) actually exceeds unity contradicting this revised analysis. This finding supports the view that a global convergence to an uncontrolled *R*_*o*_ = 4.5 is a more likely explanation than a power-law alternative arising from a non-linear force of infection in an SIR framework. To be clear, there are other causes for power-law solutions (e.g. a stochastic *β* as discussed elsewhere [31]), but those fall outside the domain of deterministic SIR approaches adopted here. Nonetheless, and as a bridge between the studies reporting power-law growth in time for *I* and the modified SIR here, a relation between *m* and *a* is sought.

The solution to equation 10 can be expressed as

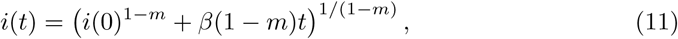

which is a power-law in *t*. For dimensionless time *γt >>* (*i*(0)^1*− m*^*/*[*R*_*o*_(1*− m*)], *i*(*t*) *∼ t*^1*/*(1*− m*)^ (*m <* 1). It directly follows that *m* = (*a −* 1)*/a*¡1 (as expected), where *a >* 1 is determined by regressing early-times log(*i*) versus log(*t*). Reported *a* for what has been termed as ‘explosive’ cases such as the US, UK Canada, Russia, among others [29] all yield an *a >* 8 (with the US *a >* 16). Such high *a* simply confirms that *m≈* 1 (and without much variations), and the early time SIR dynamics does describe reasonably those cases. For low *a* values, termed as ‘steady’, the mean *a ≈* 4.8), and thus yields an *m≈* 0.8, still not too far from unity. The shortcoming of analyzing *I*(*t*) upon *t* is that absolute figures of *I*(*t*) are sensitive to increased COVID-19 testing in time, which is considered next.

### Impact of testing ramp-up

A further explanation of early-time deviation from the SIR model (noted in several data sets here) may be time-dependent ramp-up of testing, which reveals existing infections at a rate faster than the infection spread. This hypothesis can be implemented in the SIR model considering the temporal dynamics of the testing capacity, *f*. Assuming the maximum fraction of individuals that can be tested is *f* = 1 and testing capacity grows at a rate *k*, independent of *I*, gives, *f* (*t*) = 1 *−* exp (*kt*). Therefore, the apparent number of infections, *i*_*a*_, initially grows according to the superposition of the infectious spread rate and testing capacity increase rate, i.e., exp [(*R*_0_ *−* 1 + *k*)*t*] and log (*di*_*a*_*/dt*) *∼* (*R*_0_ *−* 1 + *k*) log (*i*_*a*_). Therefore, the log-log slope will be initially greater than (*R*_0_ *−* 1) while the rate of testing increases and then converge to (*R*_0_ *−* 1) asymptotically as testing reaches steady-state. Indeed, this effect is widely observed in the global and smaller-scale data, indicating that the imprint of testing ramp-up fully dissipates (at least at the country scale) and the observed convergent slope remains a robust indicator of the early phases of virus dynamics.

### Size of the epidemic

The maximum infections *I*_*max*_ (where *dI/dt* = 0) can be derived as a function of *S*_*o*_ and *R*_*o*_ by first dividing the budgets of *dS/dt* and *dI/dt*, solving the resulting equation, and noting that *dI/dt* = 0 when *S*(*t*)*/S*_*o*_ = *γ/β* at *I*_*max*_ to yield

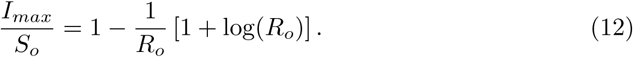

Variations of *I*_*max*_*/S*_*o*_ versus *R*_*o*_ are featured in Figure 6

**Fig 6.**
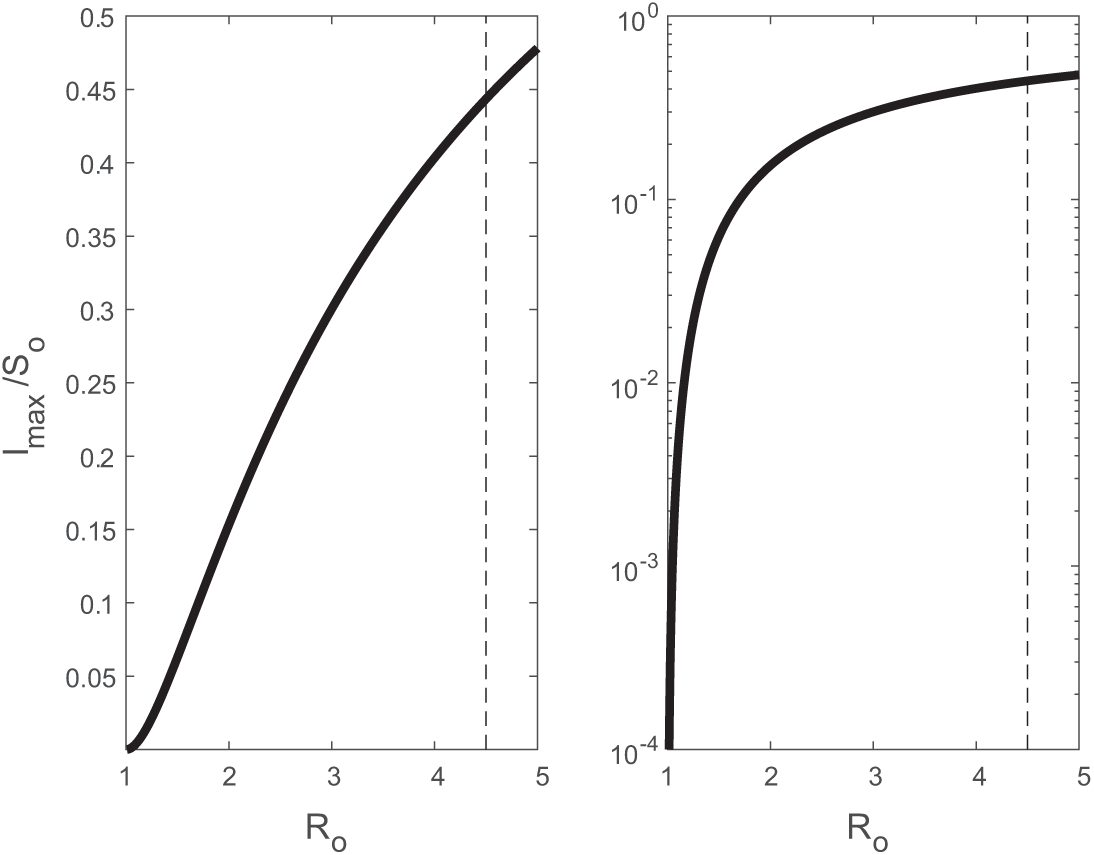
Relation between maximum infection fraction *I*_*max*_*/S*_*o*_ and *R*_*o*_.

For *R*_*o*_ = 4.5, *I*_*max*_*/S*_*o*_ = 0.44, which is much higher than values obtained for the common cold or the flu (*R*_*o*_ = 2 *−* 3) or influenza (*R*_*o*_ = 1.4 *−* 2.8). The most significant use of *R*_*o*_ is an estimate of the size of the epidemic. The total fraction of infected individuals may be inferred from 1 *− S*(*∞*)*/S*(0), where

*S*(*∞*)*/S*(0) = 1*− R*(*∞*)*/S*(0) *>* 0 because *I*(*∞*) = 0. The relation between *S*(*t*) and *R*(*t*) can be derived

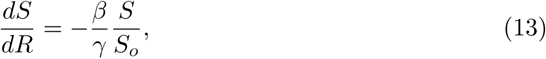

which when integrated between *t* = 0 and *t* = *∞* yields,

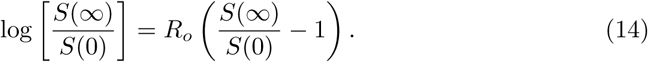

The solution requires solving a transcendental equation for *S*(*∞*)*/S*_*o*_, which can be achieved numerically. For pre-specified *R*_*o*_, the total fraction of infected individuals is shown in Figure 7. With such a high *R*_*o*_ = 4.5, some 98% of the population will be infected. When mortality is assumed to be some fraction of the total infected individuals, then the mortality fraction is *α*_*m*_[1 *− S*(*∞*)*/S*(0)].

**Fig 7.**
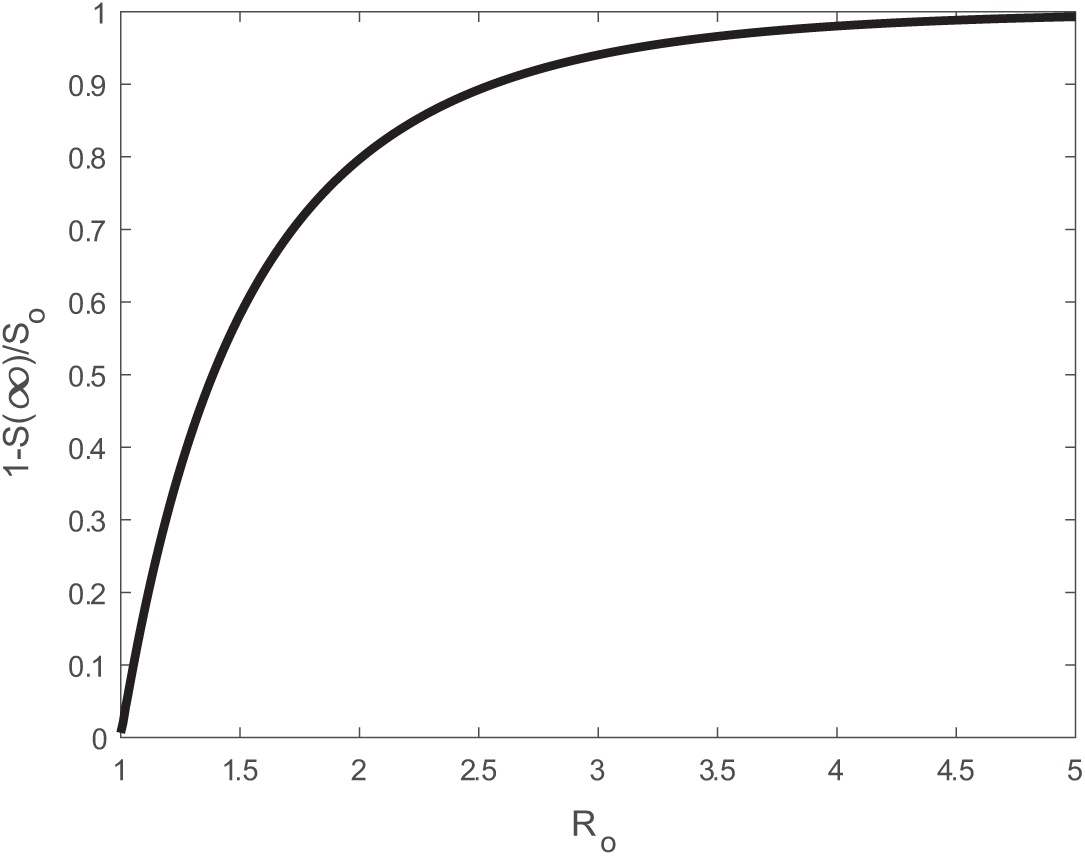
Relation between total infection fraction 1 *− S*(*∞*)*/S*_*o*_ and *R*_*o*_.

As of March 5, 2020, global mortality estimates from COVID-19 by the World Health Organization (WHO) are at *α*_*m*_ = 3.4% virtually identical to the current USA value (as of April 8, 2020; Confirmed cases = 428,901 and Mortality = 14,600). For the USA, now the epicenter of COVID-19, we ask how much *R*_*o*_ should be reduced by deliberate intervention to maintain mortality below a certain threshold size *M*_*o*_. With *S*(0) = 327M, we determine how much *R*_*o*_ should be reduced as a function of *M*_*o*_ assuming *α*_*m*_ = 3.4%. These results are featured in Figure 8 and suggest that to maintain mortality below 1 million, *R*_*o*_ *<* 1.15, a factor of 4 reduction over its uncontrolled value.

**Fig 8.**
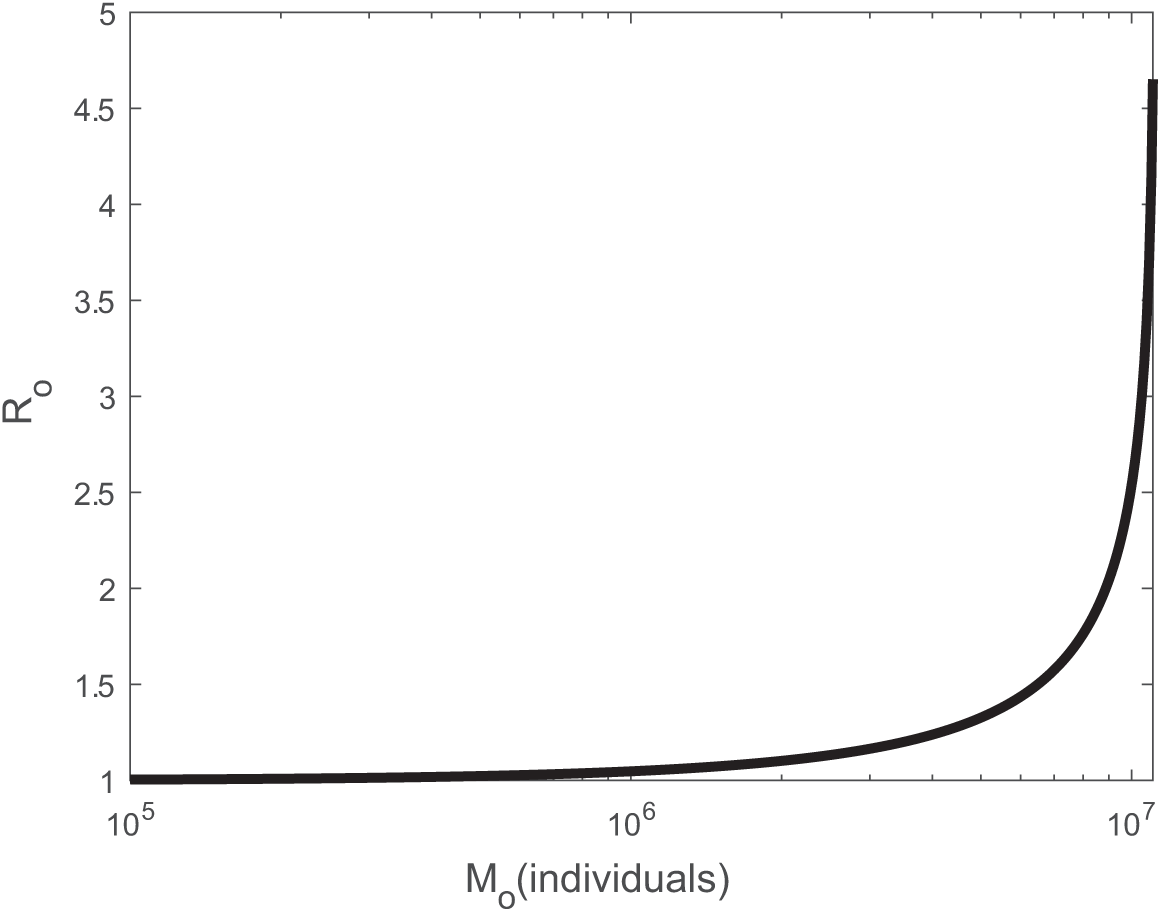
Relation between total mortality and *R*_*o*_ assuming *α*_*m*_ = 0.034 and *S*_*o*_ = 327M.

A natural extension of this exercise is to consider temporal changes in *R*_*o*_ following the logistic form in equation 6. The maximum number of infected *I*_*max*_ at time *t* and cumulative number of infections *R*(*∞*) *≈ R*(*γt ≈* 14) can be made to vary as the slope *k*_*c*_ and *t*_50_ are changed (equation 6). A larger *k*_*c*_ signifies more rapid enforcement of intervention policies and a larger *t*_50_ represents later enforcement. To provide a physical meaning for *k*_*c*_, we define 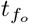 as the time after first infection at which *R*_*o*_ is *f*_*o*_% of the way through its total decline from *R*_*o,u*_ to *R*_*o,c*_. With these definitions, 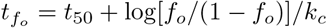. An obvious choice for *R*_*o,u*_ = 4.5, the global average when no intervention is enforced. A logical choice for *f*_*o*_ = 80% and is consistent with the point at which the logistic function enters the ‘flattening phase’. We choose *R*_*o,c*_ = 1.0 to represent the most optimistic scenario of a near-containment by intervention. For reference, the South Korea data suggests that early intervention, even when rapidly enforced shortly after the outbreak, resulted in *R*_*o*_ = 1.5 [32]. The effectiveness of interventions and any delays can now be converted to mortality and severity by varying *t*_50_ and *t*_80_ on *R*(*γt ≈* 14) and *I*_*max*_ as shown in figure 9. These figures present how *R*(*γt ≈* 14), a number connected to the cumulative number of fatalities, and *I*_*max*_, a number representing the degree to which existing resources (i.e., hospital beds will be overwhelmed), are contained for only a restricted envelope of speed and timeliness of policy enforcement. The results in Figure 9 indicate that if *R*_*o*_ *>* 2.7 within *t* = 3.5*/γ* (about 49 d here), a *>* 10% reductions relative to *S*_*o*_ in *I*_*max*_ or *R*(*∞*) are unlikely.

**Fig 9.**
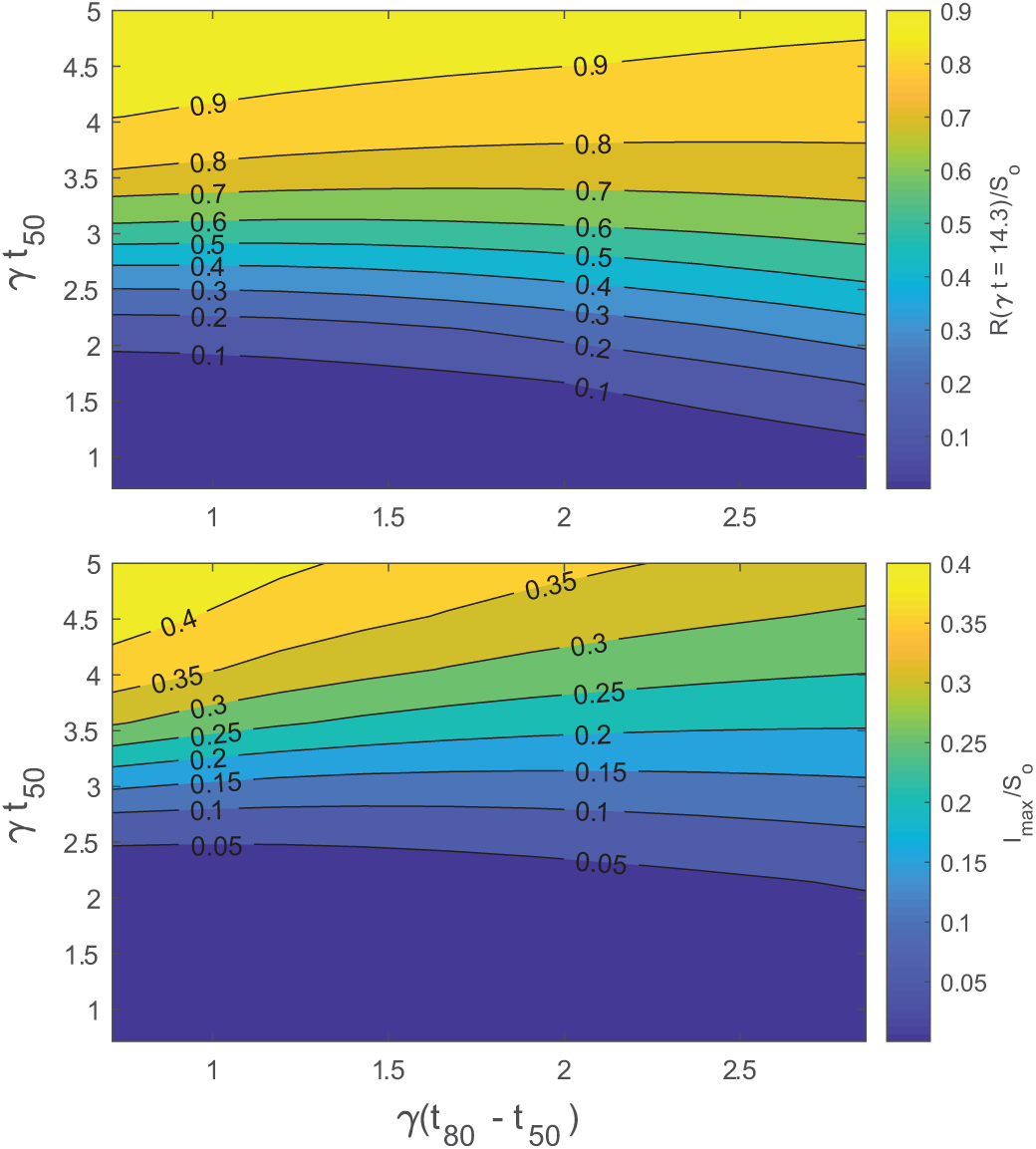
Variation in cumulative number of infected relative to *S*_*o*_ (top) and in maximum number of infected relative to *S*_*o*_ (bottom). The logistic form of *R*_*o*_ was used (equation 6). The *R*_*o*_ was set to vary from *R*_*o,c*_ = 4.5 to *R*_*o,u*_ = 1.0. *t*_50_ and *t*_80_ are the times at which *R*_*o*_ is half and 80% through the the total decline.

Back to the discussion about US mortality from COVID-19, current optimistic projections place that figure in the range of 100,000 fatalities [33]. One implication from figure 9 is that if *R*_*o*_ failed to decrease to at least 2.7 by 49 days after first infection, 8 million people are expected to die with an assumed constant mortality rate of 3.4%. For mortality to be confined to 100,000, then a reduction of *R*_*o*_ from 4.5 to 2.7 must be achieved within 17 days of first infection.

Last, it is to be noted that the fraction of individuals that must be immune (either through vaccination or recovery from prior COVID-19 infections) must exceed the herd immune threshold (HIT), which is given by

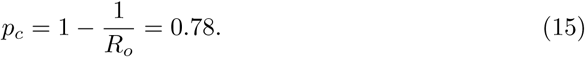

This estimate of HIT sets the limit on the immune population needed to overcome another COVID-19 pandemic (assuming a global constant *R*_*o*_ = 4.5 and no intervention). Should immunity from prior COVID-19 infections be transient, this estimate then sets the upper bound on the fraction of population that must be vaccinated and the vaccine needed in the future.

## Conclusion

The work here has shown a global convergence of *R*_*o*_ = 4.5 when no deliberate intervention was taken for COVID-19. This *R*_*o*_ was shown to describe reasonably the maximum initial exponential growth rate of COVID-19 (=(*R*_*o*_*−* 1)*γ*, where *γ* = (1*/*14)*d*^*−* 1^) in many countries that did not initiate preventive measures within *γt* = 2. The findings here further supports the growing consensus that the initial *R*_*o*_ = 2.2 estimate from Wuhan, China are low. The value of *R*_*o*_ = 4.5 is much more in line with other estimates (*R*_*o*_ = 4 *−* 6) derived from far more complex models. The critical herd immunity level that must be reached is 78% to ensure COVID-19 does not become an epidemic again. This estimate sets a maximum limit on the vaccination required. For the USA, our results show that to maintain death figures below 1*M*, the deliberate measures to be taken must reduce uncontrolled *R*_*o*_ by a factor of 4.

## Data Availability

Daily confirmed COVID-19 infections in World countries are available at https://www.ecdc.europa.eu/en/publications-data/download-todays-data-geographic-distribution-covid-19-cases-worldwide (accessed on 08/04/2020). The dataset includes population estimates for each country as obtained from the 2018 United Nations census.
Reported cases for UTLAs in the UK and provinces in Italy were obtained from https://www.gov.uk/government/publications/covid-19-track-coronavirus-cases (accessed on 09/04/2020) and https://github.com/pcm-dpc/COVID-19 (accessed on 09/04/2020), respectively. Population counts for UTLAs and Italian provinces are available at https://www.ons.gov.uk/peoplepopulationandcommunity/populationandmigration/populationestimates/datasets/populationestimatesforukenglandandwalesscotlandandnorthernireland (accessed on 04/04/2020) and http://dati.istat.it/Index.aspx?DataSetCode=DCIS_POPRES1 (accessed on 03/04/2020), respectively.

https://www.gov.uk/government/publications/covid-19-track-coronavirus-cases

https://github.com/pcm-dpc/COVID-19

https://www.ons.gov.uk/peoplepopulationandcommunity/populationandmigration/populationestimates/datasets/populationestimatesforukenglandandwalesscotlandandnorthernireland

http://dati.istat.it/Index.aspx?DataSetCode=DCIS_POPRES1

## Data availability

Daily confirmed COVID-19 infections in World countries are available at https://www.ecdc.europa.eu/en/publications-data/download-todays-data-geographic-distribution-covid-19-cases-worldwide (accessed on 08/04/2020). The dataset includes population estimates for each country as obtained from the 2018 United Nations census. Reported cases for UTLAs in the UK and provinces in Italy were obtained from https://www.gov.uk/government/publications/covid-19-track-coronavirus-cases (accessed on 09/04/2020) and https://github.com/pcm-dpc/COVID-19 (accessed on 09/04/2020), respectively. Population counts for UTLAs and Italian provinces are available at https://www.ons.gov.uk/peoplepopulationandcommunity/populationandmigration/populationestimates/datasets/populationestimatesforukenglandandwalesscotlandandnorthernireland (accessed on 04/04/2020) and http://dati.istat.it/Index.aspx?DataSetCode=DCIS_POPRES1 (accessed on 03/04/2020), respectively.

